# Rethinking immunologic risk: a retrospective cohort study of severe SARS-CoV-2 infections in individuals with congenital immunodeficiencies

**DOI:** 10.1101/2023.06.01.23290843

**Authors:** Alan A. Nguyen, Saddiq B. Habiballah, Brenna LaBere, Megan Day-Lewis, Megan Elkins, Amer Al-Musa, Anne Chu, Jennifer Jones, Ari J. Fried, Douglas McDonald, David P. Hoytema van Konijnenburg, Shira Rockowitz, Piotr Sliz, Hans C. Oettgen, Lynda C. Schneider, Andrew MacGinnitie, Lisa M. Bartnikas, Craig D. Platt, Toshiro K. Ohsumi, Janet Chou

**Author notes:** **Corresponding author:** Janet Chou, MD, Boston Children’s Hospital, 300 Longwood Avenue, Boston, MA 02115. A. A. Nguyen, S. B. Habiballah, and B. LaBere contributed equally. T. K. Ohsumi and J. Chou contributed equally. Department of Pediatrics, Faculty of Medicine, King Abdulaziz University, Jeddah, KSA (S.B.H). **Disclosures:** No authors have disclosures relevant to the content of this manuscript.

## Abstract

**Background:** Debates on the allocation of medical resources during the COVID-19 pandemic revealed the need for a better understanding of immunologic risk. Studies highlighted variable clinical outcomes of SARS-CoV-2 infections in individuals with defects in both adaptive and innate immunity, suggesting additional contributions from other factors. Notably, none of these studies controlled for variables linked with social determinants of health.

**Objective:** To determine the contributions of determinants of health to risk of hospitalization for SARS-CoV-2 infection among individuals with inborn errors of immunodeficiencies.

**Methods:** This is a retrospective, single-center cohort study of 166 individuals with inborn errors of immunity, aged two months through 69 years, who developed SARS-CoV-2 infections from March 1, 2020 through March 31, 2022. Risks of hospitalization was assessed using a multivariable logistic regression analysis.

**Results:** The risk of SARS-CoV-2-related hospitalization was associated with underrepresented racial and ethnic populations (odds ratio [OR] 5.29; confidence interval [CI], 1.76-17.0), a diagnosis of any genetically-defined immunodeficiency (OR 4.62; CI, 1.60-14.8), use of B cell depleting therapy within one year of infection (OR 6.1; CI, 1.05-38.5), obesity (OR 3.74; CI, 1.17-12.5), and neurologic disease (OR 5.38; CI, 1.61-17.8). COVID-19 vaccination was associated with reduced hospitalization risk (OR 0.52; CI, 0.31-0.81). Defective T cell function, immune-mediated organ dysfunction, and social vulnerability were not associated with increased risk of hospitalization after controlling for covariates.

**Conclusions:** The associations between race, ethnicity, and obesity with increased risk of hospitalization for SARS-CoV-2 infection indicate the importance of variables linked with social determinants of health as immunologic risk factors for individuals with inborn errors of immunity.

**Highlights:**

1. ***What is already known about this topic?*** Outcomes of SARS-CoV-2 infections in individuals with inborn errors of immunity (IEI) are highly variable. Prior studies of patients with IEI have not controlled for race or social vulnerability.
2. ***What does this article add to our knowledge***? For individuals with IEI, hospitalizations for SARS-CoV-2 were associated with race, ethnicity, obesity, and neurologic disease. Specific types of immunodeficiency, organ dysfunction, and social vulnerability were not associated with increased risk of hospitalization.
3. ***How does this study impact current management guidelines?*** Current guidelines for the management of IEIs focus on risk conferred by genetic and cellular mechanisms. This study highlights the importance of considering variables linked with social determinants of health and common comorbidities as immunologic risk factors.

## Introduction

The COVID-19 pandemic prompted dilemmas and debates regarding the allocation of medical resources.^1–3^ These debates highlighted a critical gap in medicine that remains today: the paucity of established measures of immunologic risk. This contrasts with the extensive framework of risk assessment that is the bedrock of preventative medicine. The United States Department of Health and Human Services Healthy People 2030 screening recommendations include assessments for cardiovascular disease, osteoporosis, cancer, sexually transmitted diseases, mental health, diabetes, child development, pregnancy, and sensory or communication disorders^4^. While these recommendations were developed pre-pandemic, COVID-19 has heightened the importance of screening for immunologic risk which is notably absent.

Current knowledge of immunologic risk is largely derived from clinical outcomes of primary or secondary immunodeficiencies. Primary immunodeficiencies, commonly referred to as inborn errors of immunity, consist of 485 disorders caused by defects in genes important for host immunity.^5^ Studies have identified cellular and molecular mechanisms of susceptibility to SARS-CoV-2 among the four main types of inborn errors of immunity: (1) combined immunodeficiencies with diminished T and B cell function, (2) humoral immunodeficiencies impairing B cell function, (3) defective innate immunity affecting innate immune cells or signaling, and (4) primary immune regulatory disorders with autoimmunity as the dominant feature (**Figure 1A**).^6,7^ In the U.S., national guidelines for prioritizing anti-SARS-CoV-2 therapies highlighted defective T cell function as a high risk condition due to the established role of T cells in orchestrating serologic immunity and eradicating virus-infected cells.^8^ Surprisingly, patients with severe combined immunodeficiency exhibited a spectrum of outcomes with SARS-CoV-2 infections, ranging from mild to fatal In addition to highlighting the important of innate immunity, multiple studies revealed conflicting outcomes of SARS-CoV-2 infections in individuals with other types of inborn errors of immunity,^6,9–22^ suggesting additional risk factors beyond mechanistically-defined immune defects. As these studies were predominantly comprised of adults and few included multivariate analyses, the relative contributions of age, comorbidities, and immunologic defects were not delineated. None of these studies controlled for race, Hispanic ethnicity, or social vulnerability. Thus, the relative contributions of molecular risk factors compared to socioeconomic disparities to SARS-CoV-2 infection outcomes remain unknown.

**Figure 1.**
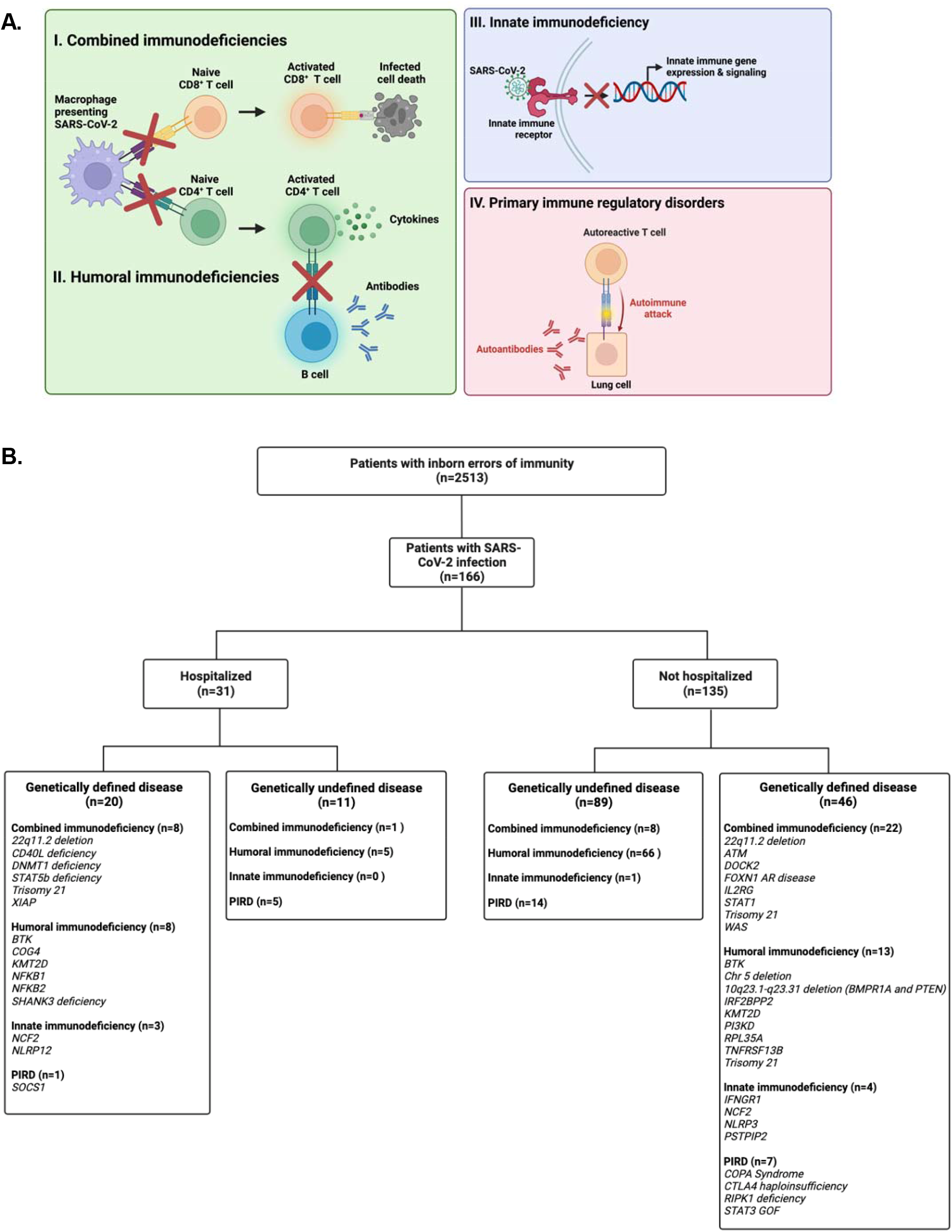
Summary of study design. **(A)** Affected pathways implicated in the four main categories of inborn errors of immunity. **(B)** Flow diagram of study participants.

We hypothesized that general risk factors and variables associated with health disparities such as race and ethnicity correlate with increased immunologic risk, even in a population with mechanistically defined immunodeficiencies. We performed a two-year retrospective cohort study of 166 patients with inborn errors of immunity, aged two months through 69 years of age. Of published studies to date, this is the largest and longest study of outcomes from SARS-CoV-2 infections in patients with inborn errors of immunity.

## Methods

### Study design and setting

This is a retrospective, single-center cohort study at Boston Children’s Hospital. As one of the nation’s largest clinical immunology programs, the Division of Immunology provides longitudinal outpatient care for patients with inborn errors of immunity ranging from the neonatal period through 75 years of age and inpatient care for individuals until the age of 35 years. Hospitalization for those older than 35 years is coordinated at adult hospitals. Patients in our cohort had a median follow-up of 9.1 years in our immunology clinic. This study was approved by the Boston Children’s Hospital Institutional Review Board.

### Study objectives, variables, and outcome

We aimed to determine factors associated with severe SARS-CoV-2 infections in patients with inborn errors of immunity, with hospitalizations as the primary outcome. Data were abstracted from the Boston Children’s Hospital electronic medical record by clinical immunologists using a standardized data collection instrument. To reduce bias inherent in a retrospective study, variables were defined prior to study initiation: age, race, ethnicity, social vulnerability index (SVI), past medical history, and history of vaccinations and therapeutics against SARS-CoV-2. Race and ethnicity were classified based on patient self-report in the electronic medical record. Patient zip codes were matched to census tracts for determination of SVI.^23^ For zip codes contained within multiple census tracts, a weighted average SVI was calculated using the American Community Survey 2014–2018 population estimates.^24^ Defective T cell function was defined as 30% of normal PHA proliferation and/or history of opportunistic infections indicative of impaired T cell function^25^. We identified patients with organ dysfunction, as defined by: (1) histological evidence of immune cell infiltration in organs, or (2) laboratory or otherwise measurable evidence of impaired organ function attributed by the patient’s primary immunologist to recurrent infections, autoimmunity, or chronic inflammation. Organ dysfunction criteria were met prior to COVID-19 infections. Obesity was defined based on CDC definitions of weight classification: in patients under 20 years of age, BMI at the 95th percentile or higher was used; in patients over 20 years of age, BMI of 30 or greater was used. Reasons for hospitalization were determined by chart review, as specified by the admission note. Respiratory insufficiency was defined as increased work of breathing, hypoxia, or respiratory failure. Hemodynamic instability was defined as unstable or low blood pressure requiring intervention. Worsening immune dysregulation was defined as evidence of active disease-specific organ dysfunction or cytopenias. Fever and neutropenia were defined as temperatures >38C and absolute neutrophil count <500 cells/microliter. Severe gastrointestinal symptoms included vomiting and/or diarrhea causing electrolyte derangements and/or hypotension requiring intervention. Clinical monitoring due to clinician concern was dependent on physician concern related to the patient’s baseline health status, diagnosis, and perceived risk in the setting of acute SARS CoV-2.

### Patients

Patients eligible for this study had (1) a diagnosis of an inborn error of immunity and (2) positive nucleic acid amplification test or antigen test for SARS-CoV-2 infection between March 15, 2020 through March 31, 2022 (**Figure 1B**). Specifically, patients with inborn errors of immunity were first identified by an ICD-10 code (**Figure E2**) used for a clinical visit between January 1, 2015 and January 1, 2020, which was verified through a medical record review by this study’s clinical immunology team. Patients with positive testing for SARS-CoV-2 were identified through review of: (1) SARS-CoV-2 testing results available at Boston Children’s Hospital, (2) referrals for SARS-CoV-2 therapeutics, (3) and medical records. Patients were not eligible if they had a secondary or acquired immunodeficiency. There were no inpatients incidentally found to have positive SARS-CoV-2 RT-PCR while hospitalized for diagnoses unrelated to COVID-19.

### Statistical analyses

We used a multivariable logistic analysis to identify potential risk factors for hospitalization due to SARS-CoV-2. Variance inflation factors were used to assess multicollinearity of potential covariates^26^. Since hospitalization for acute SARS-CoV-2 infections are relatively rare until the sixth decade of life^27^, we performed the logistic regression with penalized likelihood to reduce the bias inherent in small sample sizes^28^. As a confirmatory approach, we also identified risk factors for hospitalization using a standard logistic regression model with the same variables. There was no missing data in this data set. Statistical analyses were performed with Statistical Package for the Social Science (SPSS, version 27.0 software) and R Statistical Software (version 4.1.3).

## Results

### Patient Characteristics

Among 2513 patients with a confirmed diagnosis of an inborn error of immunity, 166 had at least one SARS-CoV-2 infection during the study period (**Figure 1B**). Thirty-one patients were hospitalized (**Table 1**), most commonly for respiratory failure (**Table 2**). Twelve patients required critical care and one died. The median age of inpatients was 16.3 years (range 0.5 – 51.7 years), compared with 14.9 years for outpatients (range 0.2 – 68.8 years). The percentage of hospitalized patients (18.7%, with a median age of 16.3 years) is higher than that of the general pediatric and adolescent population, which has been estimated to be less than 5%.^29,30^ Hospitalized patients were more likely to be male, of Black, Asian/Pacific Islander, American Indian, or Alaskan Native racial backgrounds, of Hispanic ethnicity, and living in an area with a higher social vulnerability index (**Table 1**). Of the 36 individuals who were of Black, Asian/Pacific Islander, American Indian, or Alaskan Native racial backgrounds or Hispanic ethnicity, 42.4% were hospitalized. Sixty-four patients (38%) had COVID-19 in the period after December 31, 2021 when Omicron accounted for the majority of infections in Massachusetts.

**Table 1.** Patient characteristics.

|  | **Hospitalized**<br>**n = 31** | **Outpatient**<br>**n = 135** |
| --- | --- | --- |
| **Median age, years** (range) | 16.3 (0.5 – 51.7) | 14.9 (0.2 – 68.8) |
| **Age subgroups, n (%)** |  |  |
| 0.2 – 5 years | 6 (19.4) | 18 (13.3) |
| 5 – 12 years | 8 (25.8) | 36 (26.7) |
| 12 – 18 years | 6 (19.4) | 37 (27.4) |
| 18 – 29 years | 8 (25.8) | 29 (21.5) |
| 29 – 69 years | 3 (9.6) | 15 (11.1) |
| **Biologic sex, n (%)** |  |  |
| Male | 18 (58.1) | 73 (54.1) |
| Female | 13 (41.9) | 62 (45.9) |
| **Self-reported race, n (%)** |  |  |
| Asian/Pacific Islander | 2 (6.5) | 2 (1.5) |
| Black | 3 (9.7) | 6 (4.4) |
| American Indian or Alaska Native | 1 (3.2) | 0 |
| White | 17 (54.8) | 116 (86.0) |
| Other<sup>a</sup> | 8 (25.8) | 11 (8.1) |
| **Self-reported ethnicity, n (%)** |  |  |
| Hispanic | 8 (25.8) | 8 (5.9) |
| Non-Hispanic | 23 (74.2) | 127 (94.1) |
| **Median Social vulnerability index** (range) | 0.41 (0.10 – 0.88) | 0.28 (0.018 – 0.97) |
| **Subtypes of inborn errors of immunity, n (%)** |  |  |
| Humoral immune defects | 12 (37.5) | 83 (61.5) |
| Combined immune deficiency | 11 (35.5) | 30 (22.2) |
| Innate immune defect | 3 (9.7) | 4 (3.0) |
| Primary immune dysregulatory disorder | 5 (16.1) | 18 (13.3) |
| **Clinical features and comorbidities, n (%)** |  |  |
| Genetically-defined inborn error of immunity | 20 (64.5) | 46 (34.1) |
| End-organ dysfunction | 14 (45.2) | 46 (34.1) |
| Rituximab treatment at time of infection | 5 (16.1) | 6 (4.4) |
| Obesity | 10 (32.3) | 22 (16.3) |
| Neurologic disease | 8 (25.8) | 16 (11.9) |
| Chronic renal disease | 4 (12.9) | 3 (2.2) |
| Cardiovascular disease | 4 (12.9) | 13 (9.6) |
| History of organ or stem cell transplantation | 2 (6.5) | 7 (5.2) |
| Malignancy | 1 (3.2) | 2 (1.5) |
| Chronic liver disease | 1 (3.2) | 6 (4.4) |
| Trisomy 21 | 1 (3.2) | 3 (2.2) |
| **COVID-19-related therapeutics n (%)** |  |  |
| Number of COVID-19 vaccine doses<sup>b</sup> |  |  |
| 0 | 24 (77.4) | 77 (57.0) |
| 1 | 3 (9.7) | 1 (0.7) |
| 2 | 2 (6.5) | 31 (23.0) |
| 3 | 2 (6.5) | 26 (19.3) |
| Monoclonal antibodies against SARS-CoV-2<sup>c</sup> | 4 (12.9) | 30 (23.0) |

a. For self-reported race, “other” denotes those patients who selected “other” as their racial background
b. All COVID-19 vaccines were given prior to April 2022, which excluded the bivalent formulations
c. Monoclonal antibodies included sotrovimab, bamlanivimab, casirivimab, imdevimab, and tixagevimab/cilgavimab

**Table 2.** Reasons for hospitalization*.

| **Cause** | **Number (%)** |
| --- | --- |
| Respiratory instability | 13 (42) |
| Hemodynamic instability | 3 (10) |
| Worsening immune dysregulation | 4 (13) |
| Fever and neutropenia | 2 (6) |
| Severe gastrointestinal symptoms | 4 (13) |
| Multisystem Inflammatory Syndrome in Children | 3 (10) |
| Clinical monitoring due to clinician concern | 2 (6) |

\*See Methods for information about category designation

Humoral immunodeficiencies were the most common type of inborn error of immunity, followed by combined immunodeficiencies (**Table 1**). Among inpatients, 64.5% had a genetically defined immune disorder attributable to a genetic variant confirmed by clinical-grade DNA sequencing, compared to 34.1% of outpatients (**Table 1**). Organ dysfunction caused by recurrent infections and/or autoimmunity is a complication of inborn errors of immunity that may be treated with B cell-depleting therapies.^31^ Inpatients were more likely to have organ dysfunction and B cell depleting therapies within one year prior to infection, compared to outpatients. Among comorbidities known to be associated with severe COVID-19,^32^ obesity, neurologic disease, chronic renal disease, and cardiovascular disease were the most common and occurred more frequently in inpatients than outpatients (**Table 1**).

### Use of COVID-19 medical interventions

FDA emergency authorization for COVID-19 vaccines was granted in December 2020, but distribution was hampered by limitations in supply, logistics, and public trust.^33^ The majority of inpatients (74.2%) and outpatients (57.0%) were infected with SARS-CoV-2 before COVID-19 vaccination. Additionally, 35 patients received post-exposure treatment with monoclonal antibodies SARS-CoV-2 as they were asymptomatic but tested positive, in accordance with FDA recommendations, three of whom required subsequent hospitalization due to worsening disease.

### Risk factors associated with hospitalization among patients with inborn errors of immunity

Among demographic factors, Black, Asian/Pacific Islander, American Indian, or Alaskan Native racial background and/or Hispanic ethnicity was significantly associated with an increased risk of hospitalization (OR 5.29; CI, 1.75-17.0), while social vulnerability index was not. Older age was associated with a marginally increased risk of hospitalization (OR 1.05; CI, 1.01-1.10). Hospitalization was also associated with the following clinical features relevant to inborn errors of immunity: a history of a genetically defined immunodeficiency (OR 4.62; CI, 1.60-14.8) and treatment with B cell depleting agents within one year of infection (OR 6.10; CI, 1.05-38.5). Notably, neither defective T cell function nor a history of immune-associated organ dysfunction was associated with an increased risk of hospitalization. Among the comorbidities associated with severe COVID-19 in the general population, obesity (OR 3.74; CI, 1.17 to 12.5) and a history of neurologic disease (OR 5.38; CI, 1.61 to 17.8), were associated with increased risk of hospitalization. Infection after December 31, 2021, at which point the Omicron variant accounted for over 95% of SARS-CoV-2 infections in Massachusetts, was not associated with increased hospitalizations. With respect to SARS-CoV-2 interventions, an increased number of COVID-19 vaccination doses was associated with reduced hospitalization risk (OR 0.52; CI, 0.31-0.81), while treatment with SARS-CoV-2 monoclonal antibody was not statistically significant. A standard logistic regression model identified the same risk factors with similar adjusted ORs (**Figure 2** and **Figure E1**).

**Figure 2.**
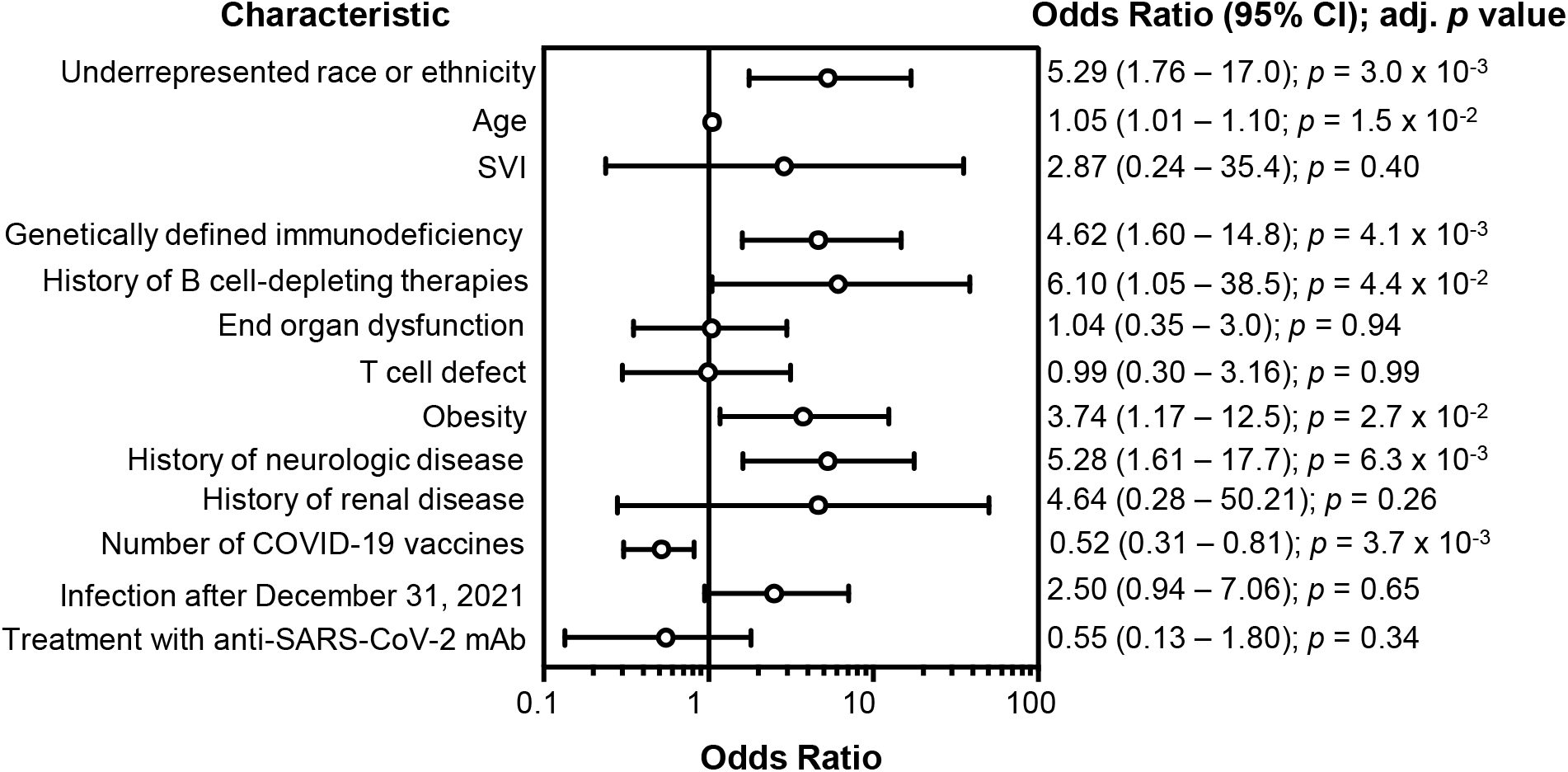
Adjusted risk of SARS-CoV-2-related hospitalization in individuals with inborn errors of immunity. Odds ratios and 95% confidence intervals (CI) for each respective determinant of health, as calculated by logistic regression with covariate analysis. After December 31, 2021, the Omicron variant accounted for at least 95% of SARS-CoV-2 infections.

## Discussion

Although inborn errors of immunity are rare diseases, they serve as real-world disease models that enable delineation of specific pathways contributing to immunologic risk. While rare disease research focuses on disease-specific principles toward a goal of personalized medicine, common risk factors for disease can be overlooked. Prior studies have not assessed the contributions of race, ethnicity, or social vulnerability to outcomes from SARS-CoV-2 infection in individuals with inborn errors of immunity.^6,9–21^ Our study reveals the importance of contextualizing host immunity within common determinants of health, which are significant contributors to outcomes from SARS-CoV-2 infections.

We found that obesity was associated with increased odds of hospitalization, concordant with SARS-CoV-2 outcomes in the general pediatric and adult populations.^34,35^ Guidelines for clinical management of inborn errors of immunity highlight low body weight as a predictor of immunologic compromise.^21,25^ However, the prevalence of obesity in pediatric and adult patients with inborn errors of immunity is now comparable to that of the general population.^36^ During the H1N1 influenza pandemic, obesity was also associated with increased morbidity and mortality in the general population.^37^ Adipose tissue can serve as a reservoir for viral replication, the priming of inflammatory myeloid and lymphoid cells, and a source of inflammatory cytokines and adipokines.^38^ Future studies are needed to determine how obesity influences clinical outcomes other than SARS-CoV-2 infections in patients with inborn errors of immunity.

Here, we show that underrepresented racial and ethnic groups, but not SVI, are associated with increased odds of hospitalization. The lack of association between SVI and SARS-CoV-2-related hospitalizations in our study may arise from the availability of medical coverage for children and adolescents living in Massachusetts, which is the highest in the U.S.: only 1.2% of children in Massachusetts are uninsured, compared to a national average of 5.2%.^39^ Approximately one third of children in Massachusetts receive healthcare through the state’s Medicaid program providing school-based medical care.^39^ Additionally, this study was conducted at a tertiary medical center providing immunology subspecialty care. However, the persistent disparities in healthcare are underscored by the 5.2-fold odds of hospitalization associated with Black, Black, Asian/Pacific Islander, American Indian, or Alaskan Native race and/or Hispanic ethnicity in this study. This is concordant with studies in the general pediatric population showing increased hospitalizations and deaths from SARS-CoV-2 infections among non-Hispanic Black or Hispanic children compared to White children.^40,41^

Race and ethnicity have inextricably been linked with disadvantages in SDOH for minority groups when pertaining to specific conditions, including cardiovascular disease^42^ and diabetes.^43^ With regards to COVID-19, on a county level in the US there is association between adverse SDOH factors, percent Black residents, and mortality rate. However, in counties with below average adverse SDOH factors there was no association between percent Black residents and mortality, implying that racial disparities are driven by social constructs.^44^ Specific to the pediatric population, there are significant racial and ethnic differences across all categories of SDOH, including economic stability, education access and quality, health care access and quality, neighborhood and built environment, and social and community context.^45^ Interestingly, within our cohort race and ethnicity were associated with hospitalizations independent of SVI, indicating that additional studies are needed to delineate how underlying inequities and structural racism affect other outcomes in patients with inborn errors of immunity.

Our findings identify a potential role for genetic sequencing in assessing immunologic risk. The heterogenous clinical phenotypes of inborn errors of immunity complicate the diagnosis and management of these disorders. Defining the underlying genetic etiology improves understanding of patient phenotypes can guide directed therapies.^1^ Despite the declining costs of genetic sequencing, genetic tests are not routinely covered by most insurance companie.^46^ Genetic diagnoses in our cohort were made feasible by an institutional program that enables clinical whole exome sequencing at no cost to patients.^47^ In our study, patients with a genetically defined immunodeficiency had 5.3-fold greater odds of hospitalization. Within this group, there was no significant predilection among types of genetic defects: there were similar numbers of hospitalized patients with combined defects and humoral defects, with fewer patients with innate defects or PIRD (**Figure E2**). The reasons for hospitalization indicate that our findings were not attributable to clinical monitoring of patients with genetic diagnoses (**Table 2**). Rather, we posit that individuals with genetically confirmed immunodeficiencies have more severe immunologic disease and thus, a higher risk of severe SARS-CoV-2 infections. This hypothesis is supported by evidence showing that the likelihood of a genetic diagnosis is higher in patients with severe clinical presentations.^21^ Disease severity was controlled in our analysis using organ dysfunction as a surrogate. Notably, organ dysfunction – a standard measure of disease severity for disorders of immunity^31^ – was not associated with increased hospitalization risk in our cohort, even though it was present in 45.2% of inpatients and 34.1% of outpatients. Thus, genetic diagnoses may enable better identification of individuals with increased immunologic risk. Identification of genetic disorders will be made possible only through equitable access to genetic testing, a goal that remains elusive in the U.S. due to gaps in clinician knowledge, limited and inconsistent insurance coverage of genetic tests, and the scarcity of community-based or patient-centered educational resources in genomic medicine.^48^

Although hospitalization was most frequent in those with innate immune defects or combined immunodeficiencies, we did not find a significant association of defective T cell immunity with hospitalization when controlling for covariates. This may be due, at least in part, to national newborn screening programs for severe combined immunodeficiency that enable early diagnosis and treatment.^49^ Thus, most children with the most severe combined immunodeficiencies will have undergone hematopoietic stem cell transplantation before entry into daycare and school settings with potentially high levels of viral transmission. We found that increasing numbers of COVID-19 vaccinations were associated with a significantly reduced risk of hospitalization, further providing evidence that the school-aged and older individuals in this cohort had partial retention of immune function. Similarly, the largest study of mRNA vaccine efficacy in individuals with congenital immunodeficiencies found that the majority of patients with combined immunodeficiencies mounted an intact vaccine-specific T cell response.^50^ Although impaired T cell function is a commonly used measure of susceptibility to viral infections, our study highlights the limitations of applying this principle without acknowledgement of other determinants for health. Genetic diagnoses, obesity, comorbid conditions, and race modulate the immunologic risk often attributed solely to the mechanisms underlying congenital immunodeficiencies.

By using a multivariate regression model, we adjusted for variations in SARS-CoV-2 directed interventions as some patients received vaccines and monoclonal antibodies while others did not. These differences were largely due to the availability of these agents during time of infection and governmental recommendations regarding eligibility. Similar models have been used to account for confounding factors in COVID-19 studies.^51,52^ Furthermore, the single center design of our study minimized variability in clinical practice. Patients with primary immunodeficiencies and SARS-CoV-2 infection were managed using institution-approved guidelines and algorithms to standardize modalities of care.

Our study has limitations. This is a single center, retrospective study and inborn errors of immunity are rare diseases. It is infeasible to definitively determine the total number of patients with SARS-CoV-2 infections because SARS-CoV-2 test availability was limited at the start of the pandemic, patients may not report all positive home antigen tests to healthcare providers, and asymptomatic SARS-CoV-2 infections are underreported. Given the limitations inherent in studies of rare diseases with small sample sizes, our study sought to determine the contributions of common risk factors that are often overlooked in rare diseases due to the “N of 1” approach typical of personalized medicine.

The morbidity and mortality caused by the COVID-19 pandemic transformed immunologic risk from an abstract concept into a real-world axis that influences daily decisions about school, work, and travel. The findings of this study indicate the importance of considering factors beyond molecular mechanisms as measures of immunologic risk and suggest that this risk can be modified by initiatives pertaining to obesity and healthcare disparities.

## Data Availability

All data produced in the present study are available upon reasonable request to the authors.

## Abbreviations

CI: confidence interval
COVID-19: Coronavirus disease 2019
IEI: inborn errors of immunity
OR: odds ratio
SARS-CoV-2: Severe acute respiratory syndrome coronavirus 2
SDOH: social determinants of health
SVI: social vulnerability index

## Acknowledgements

This study was supported by the National Institutes of Health (T32AI007512 to H.C.O., A.A.N., B.L., D.H.; K23AI143962 to L.M.B., R01DK130465 and R01AI139633-04S1 to J.C.), the Wallace Family Fund and Perkins Fund (to J.C.), and the Department of Pediatrics at King Abdulaziz University, Jeddah, KSA (to S.B.H.).

## Figure Legends

**Figure E1. Secondary analysis of adjusted risk of SARS-CoV-2-related hospitalization in individuals with inborn errors of immunity.** Odds ratios and 95% confidence intervals (CI) derived from a standard logistic regression analysis model, after adjusting for all covariates listed. After December 31, 2021, the Omicron variant (the first SARS-CoV-2 variant to have immune escape) accounted for at least 95% of SARS-CoV-2 infections.

**Figure S2. Severity of SARS-CoV-2 infection in patients with genetically defined inborn errors of immunity.** Monogenic inborn errors of immunity separated by severity of disease and categorized by phenotypic classification.

