## Supplemental Figure E1 and E2 for "Rethinking immunologic risk: a retrospective cohort study of severe SARS-CoV-2 infections in individuals with congenital immunodeficiencies"

### Slide 1
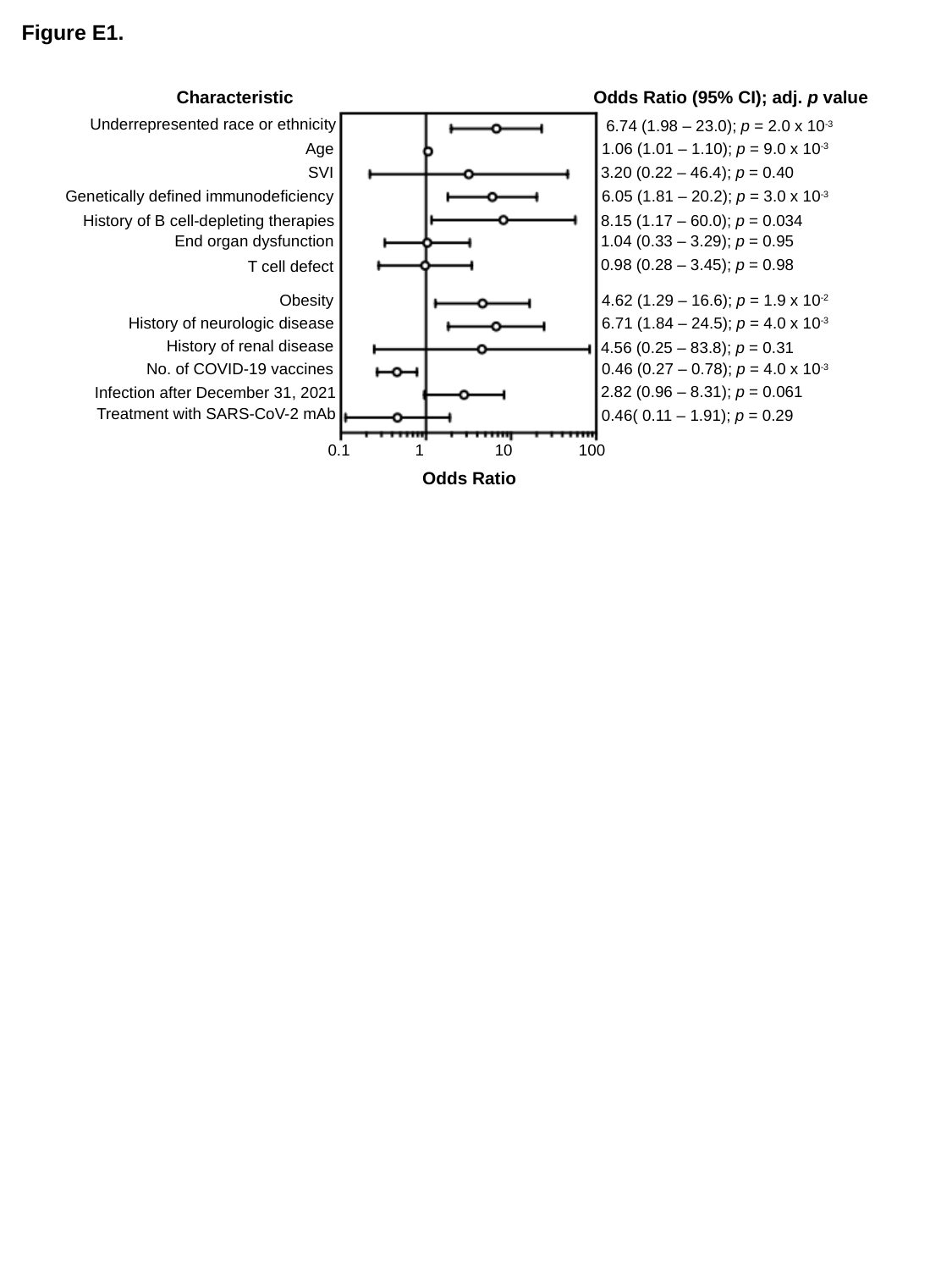

Figure E1.
Characteristic
Odds Ratio (95% CI); adj. p value
Underrepresented race or ethnicity
6.74 (1.98 – 23.0); p = 2.0 x 10^-3^
Age
1.06 (1.01 – 1.10); p = 9.0 x 10^-3^
SVI
3.20 (0.22 – 46.4); p = 0.40
Genetically defined immunodeficiency
6.05 (1.81 – 20.2); p = 3.0 x 10^-3^
History of B cell-depleting therapies
8.15 (1.17 – 60.0); p = 0.034
End organ dysfunction
1.04 (0.33 – 3.29); p = 0.95
0.98 (0.28 – 3.45); p = 0.98
T cell defect
Obesity
4.62 (1.29 – 16.6); p = 1.9 x 10^-2^
History of neurologic disease
6.71 (1.84 – 24.5); p = 4.0 x 10^-3^
History of renal disease
4.56 (0.25 – 83.8); p = 0.31
No. of COVID-19 vaccines
0.46 (0.27 – 0.78); p = 4.0 x 10^-3^
2.82 (0.96 – 8.31); p = 0.061
Infection after December 31, 2021
Treatment with SARS-CoV-2 mAb
0.46( 0.11 – 1.91); p = 0.29
0.1
1
10
100
Odds Ratio

### Slide 2
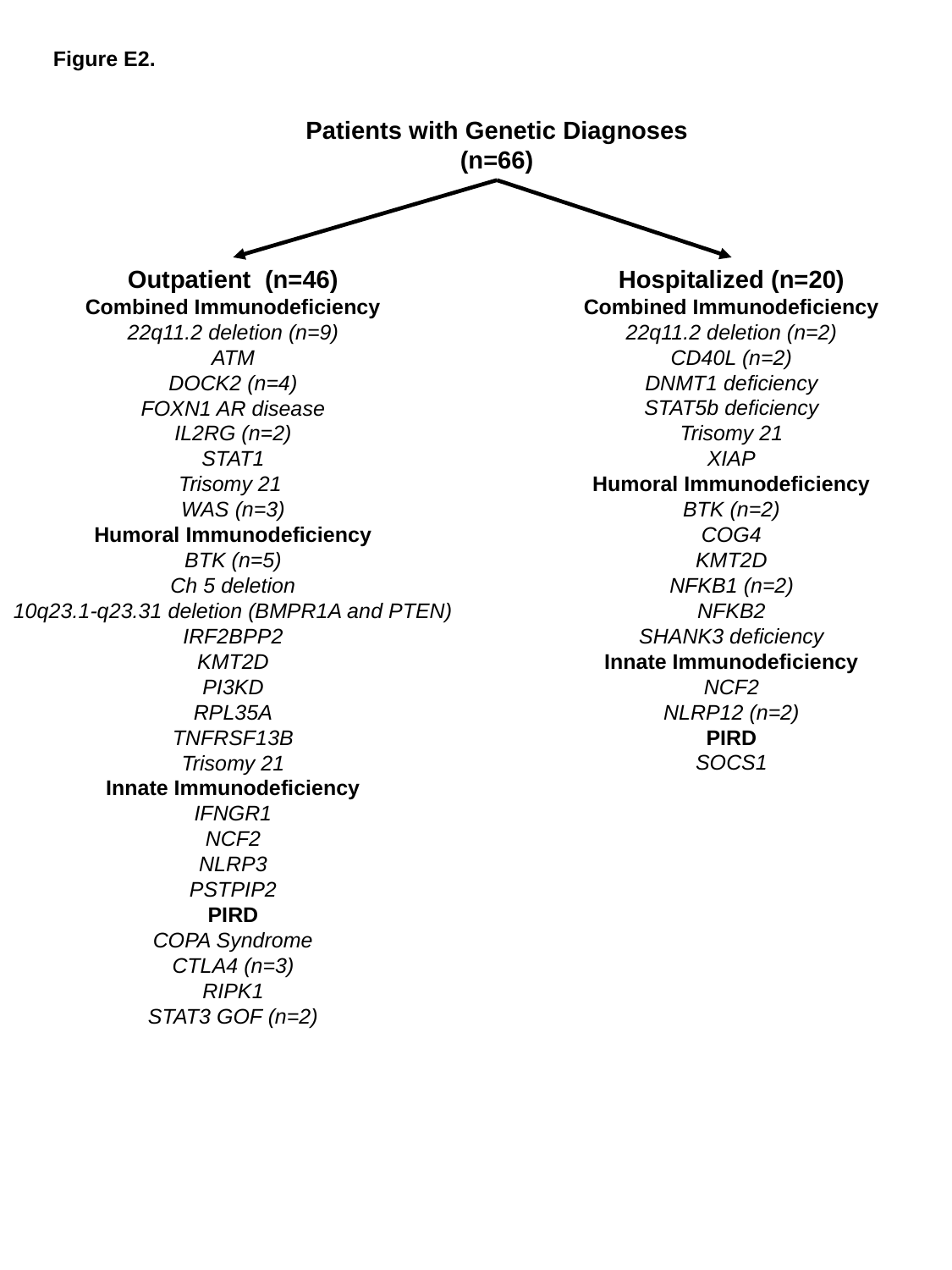

Figure E2.
Patients with Genetic Diagnoses (n=66)
Outpatient (n=46)
Combined Immunodeficiency
22q11.2 deletion (n=9)
ATM
DOCK2 (n=4)
FOXN1 AR disease
IL2RG (n=2)
STAT1
Trisomy 21
WAS (n=3)
Humoral Immunodeficiency
BTK (n=5)
Ch 5 deletion
10q23.1-q23.31 deletion (BMPR1A and PTEN)
IRF2BPP2
KMT2D
PI3KD
RPL35A
TNFRSF13B
Trisomy 21
Innate Immunodeficiency
IFNGR1
NCF2
NLRP3
PSTPIP2
PIRD
COPA Syndrome
CTLA4 (n=3)
RIPK1
STAT3 GOF (n=2)
Hospitalized (n=20)
Combined Immunodeficiency
22q11.2 deletion (n=2)
CD40L (n=2)
DNMT1 deficiency
STAT5b deficiency
Trisomy 21
XIAP
Humoral Immunodeficiency
BTK (n=2)
COG4
KMT2D
NFKB1 (n=2)
NFKB2
SHANK3 deficiency
Innate Immunodeficiency
NCF2
NLRP12 (n=2)
PIRD
SOCS1
